# Mindful and intuitive eating: A bibliometric analysis of research trends from 2004 to 2024

**DOI:** 10.1101/2025.04.29.25326674

**Authors:** Mengying Zhang, Angeliki Bogossian, Emma Stanmore, Trudi Edginton, Siobhán O’Connor

## Abstract

**Introduction:** Excessive body weight and problematic eating behaviours, such as emotional and binge eating, have become major public health concerns. Mindful eating and intuitive eating offer alternative approaches to traditional dieting by encouraging individuals to focus on internal hunger and satiety cues. This study aims to analyse research trends and metrics related to mindful eating and intuitive eating publications over the past twenty years.

**Methods:** We used Scopus to identify mindful and intuitive eating related studies from 2004 to 2024. VOSviewer and Bibliometrix were used to extract relevant data and run the bibliometric analysis.

**Results:** A total of 1922 articles and reviews published in English were identified. After screening titles and abstract for relevance, 1064 documents were left for analysis. The number of studies on mindful and intuitive eating increased most years, with 2023 (n=143, 13.44%) and 2024 (n=146, 13.72%) producing the most articles, and a total citation count of 32,245 over the twenty-year period. High-income countries such as the United States (n=497), United Kingdom (n=131), and Canada (n=88) produced the most scientific articles. Leading researchers were Mantzios, M. (n=26) and Tylka, T. L. (n=25). The most cited articles focused on mindfulness or meditation-based therapies in managing psychological stress and the influence of taste on food choices. Keyword and trend analyses highlight emerging research areas such as ‘*mindfulness*’ and ‘*intuitive eating*’.

**Conclusion:** This bibliometric analysis provides valuable insights to the research trends in the mindful and intuitive eating fields. Strengthening interdisciplinary research and expanding collaborations between countries and authors could enhance the research impact in these fields as well as exploring emerging topics such as body perceptions (e.g., body image, body positivity, body dissatisfaction).

## 1. INTRODUCTION

Eating behaviour is used to respond to hunger to regulate internal homeostasis but problematic eating behaviours such as emotional eating or binge eating can lead to excessive food intake [1, 2]. Problematic eating behaviour can stem from a number of biological, individual, and social factors including negative emotions like stress, low self-esteem, bullying and peer pressure, body dissatisfaction and dieting, poverty and food insecurity, and the availability of low-cost high-calorie food and ultra-processed food. This can lead to increased body weight and obesity along with other physical and mental health issues. For example, a systematic review found that ultra-processed food consumption was associated with a range of non-communicable diseases, including abdominal obesity, metabolic syndrome, and depression in adults [3].

Obesity is a global public health problem with the World Health Organization reporting 1 in 8 people in the world live with obesity, approximately 890 million adults and 160 million children and adolescents [4]. At the individual level, being overweight is a risk factor for several chronic illnesses including type 2 diabetes, cancer, osteoarthritis, liver, renal, respiratory and cardiovascular diseases among others. These conditions can further increase the risk of morbidities and mortality [5, 6]. The economic impact of being overweight or obese is also substantial with global estimates that it will cost $3 trillion per year by 2030 [4].

Traditional weight management methods for tackling unhealthy eating habits such as restricting calorie intake or limiting certain types of food may be effective in the short-term but can have negative effects on both physical and mental health in the long-term [7, 8]. For example, the ketogenic diet requires strict control of carbohydrate intake to promote weight loss [9]. While this method can effectively reduce weight in the short-term, long-term low-carbohydrate diets may lead to deficiencies in vitamins and minerals and affect cardiovascular health. Additionally, prolonged restriction of carbohydrates can affect serotonin levels in the brain, potentially leading to depression and increased anxiety [10].

Alternative strategies for addressing unhealthy eating habits have emerged such as mindful eating (ME) and intuitive eating (IE). Although there is no universally accepted definition of ME, it has been characterised by in the moment attention to eating, along with non-judgmental awareness of sensations (both physical and emotional) when eating [11] and a compassionate understanding of the neurobiology of craving and overeating [12]. The focus on paying attention to hunger and satiety signals may foster a healthier relationship with food and reduce the risk of binge eating [13]. Similarly, IE as an adaptive eating style emphasises reconnecting with natural physiological signals and satiety cues, and advocates for giving up any dietary restrictions or rules about healthy or unhealthy eating [14]. It may help individuals decrease excessive focus on weight, emphasising the importance of respecting the body’s physiological needs for food, rather than relying on external cues such as emotions, food availability etc. to eat [13, 15]. A number of scientific reviews examining the effectiveness of mindfulness, ME, and IE on eating behaviours have been conducted over the last decade, with some showing that mindfulness-based approaches reduced problematic eating patterns although their effects on weight loss were mixed [16, 17]. Furthermore, there is limited evidence that ME and IE based programs can influence dietary intake (i.e., energy intake or diet quality) in adults with no history of eating disorders [18]. Given the growing volume of literature on ME and IE, a historical analysis of the contribution these approaches make to science on weight management would be helpful to highlight the major trends and knowledge gaps to support future research and practice.

### 1.1 Bibliometrics

Bibliometric analysis offers a quantitative approach to analysing a body of literature within a particular scientific field, allowing an exploration of key concepts and research trends in a given topic [19]. It usually encompasses a review of scholarly literature, the analysis of citation and publication patterns, and the visualisation of connections between different areas of research. For instance, bibliometrics has been used in the field of weight management to diabetes mellitus and bariatric surgery [20], global obesity research [21], as well as sarcopenic obesity [22] among others. They revealed important insights including notable publication trends, seminal research studies in these fields, the most prolific authors, research collaborations and disciplinary contributions, and the growth of certain research trends over time, enabling knowledge gaps to be identified and areas for further research to be recommended. However, no such analysis has been undertaken on ME and IE related publications. This would help uncover important scientific trends in this area that could help advance weight management practice by professionals and patients and identify areas for further research. Hence, the aim of this bibliometric study is to quantify, describe, and compare published research on ME and IE over the past 20 years.

## 2. MATERIALS AND METHODS

### 2.1 Search strategy and data collection

The Scopus database is widely recognised as a reliable and comprehensive source of academic resources that span a wide range of influential areas in relation to health, life, physical and social sciences, and the humanities [23, 24]. Due to its broad coverage, it has been used to conduct bibliometric analysis in the field of obesity [25, 26]. For this study, data from Scopus were retrieved, covering publications from 2004 to 2024. Two search terms were combined: (1) mindful* or intuit* AND (2) eat* to identify relevant studies, with the search conducted in August 2024. Language restrictions were set to English and document types specified as ‘Article’ and ‘Reviews’, resulting in a total of 1,922 articles. Bibliometric data were exported in .CSV and .RIS format, including citation information, bibliographical information, abstract and keywords, and cited references. Covidence (www.covidence.org) was used to screen titles and abstracts to remove irrelevant studies.

### 2.2 Data analysis and visualisations

Two tools were used to produce the bibliometric analysis: 1) Bibliometrix (https://www.bibliometrix.org/, R studio was downloaded to facilitate analysis), and 2) VOSviewer 1.6.20 (https://www.vosviewer.com/). Bibliometrix is an R package that enables visualisation and network analysis, helping users explore research trends, collaborations, and impact within a scientific field [27]. VOSviewer is free visualisation software that facilitates a range of bibliometric analyses including co-authorship analysis, co-occurrence analysis, citation analysis, bibliographic coupling analysis, and co-citation analysis [28]. No ethical approval was required for this study as all data were sourced from a public database for secondary analysis.

## 3. RESULTS

After screening titles and abstract, a total of 1,064 studies were found over the 20-year period, with the first published studies referring to meditation, mindfulness or intuitive eating appearing in 1997 [29] and 1998 [30]. The number of studies increased in most years with 2023 (n=143, 13.44%) and 2024 (n=146, 13.72%) producing the highest volumes of articles (Table 1). From 1997 to 2024, the citation count increased from 75 citations in 1997 to a peak of 3676 citations in 2013, a total of 32245 citations for the whole period. In 2022 and 2024, there was a downward trend in total citations, from 1484 to 916 and citations per publication from 4.60 to 2.84. This may reflect the length of time it can take for research studies to be cited and citation counts to accrue in a given field [31]. The average citation per publication ranged from 0.10 in 1998 to 11.40 in 2011.

**Table 1.**
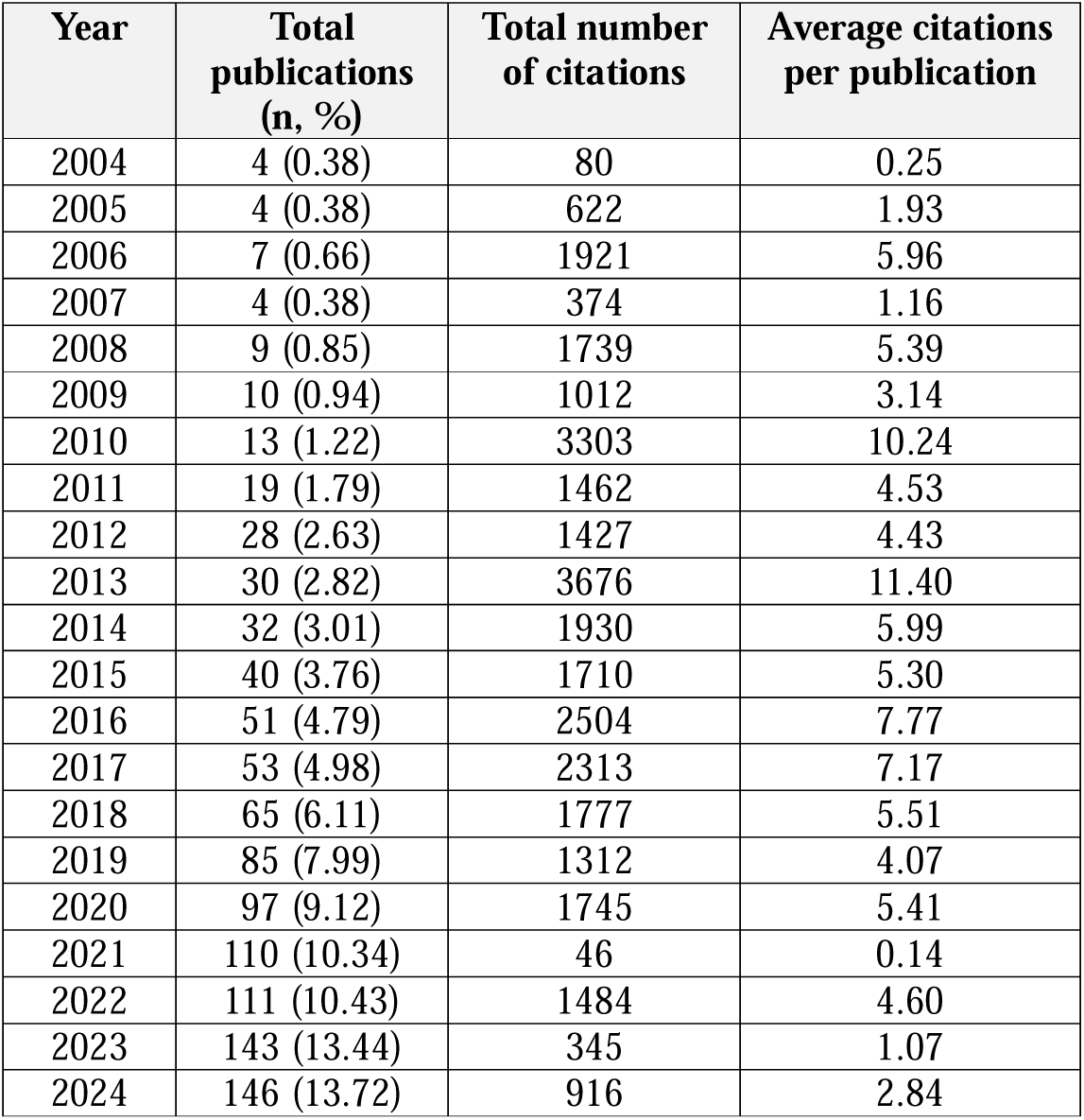
Publication metrics in ME or IE research from 1997 to 2024.

### 3.1 Country analysis

The analysis of country production and citation counts highlights the dominant role of the United States (U.S.) across multiple metrics in the ME and IE research field. In terms of corresponding author contributions, the U.S. was the top ranked country, indicating a robust domestic research output (n=376) and extensive international collaborations, with single country publication (SCP) at 342 and multiple country publication (MCP) at 34 (Figure 1). This is supported by the “Most Cited Countries” data (Figure 2), where the U.S. stands out with the highest number of citations (n=16,361), reflecting the widespread influence and recognition of this country’s research in the field. The U.K. ranks second and shows a substantial number of outputs (n=93) and citations (n=2318), emphasising its role in international research networks. Canada, ranks third in corresponding author’s number of articles (n=58), demonstrating its influence and active participation in ME and IE research. High-income countries such as Australia, the Netherlands, Germany, Portugal, New Zealand, Italy, and France consistently appear in the top ranks for both article production and article citations. Meanwhile, emerging countries such as China and Turkey are making notable strides, indicating their increasing involvement in this global research community (Table 2).

**Figure 1.**
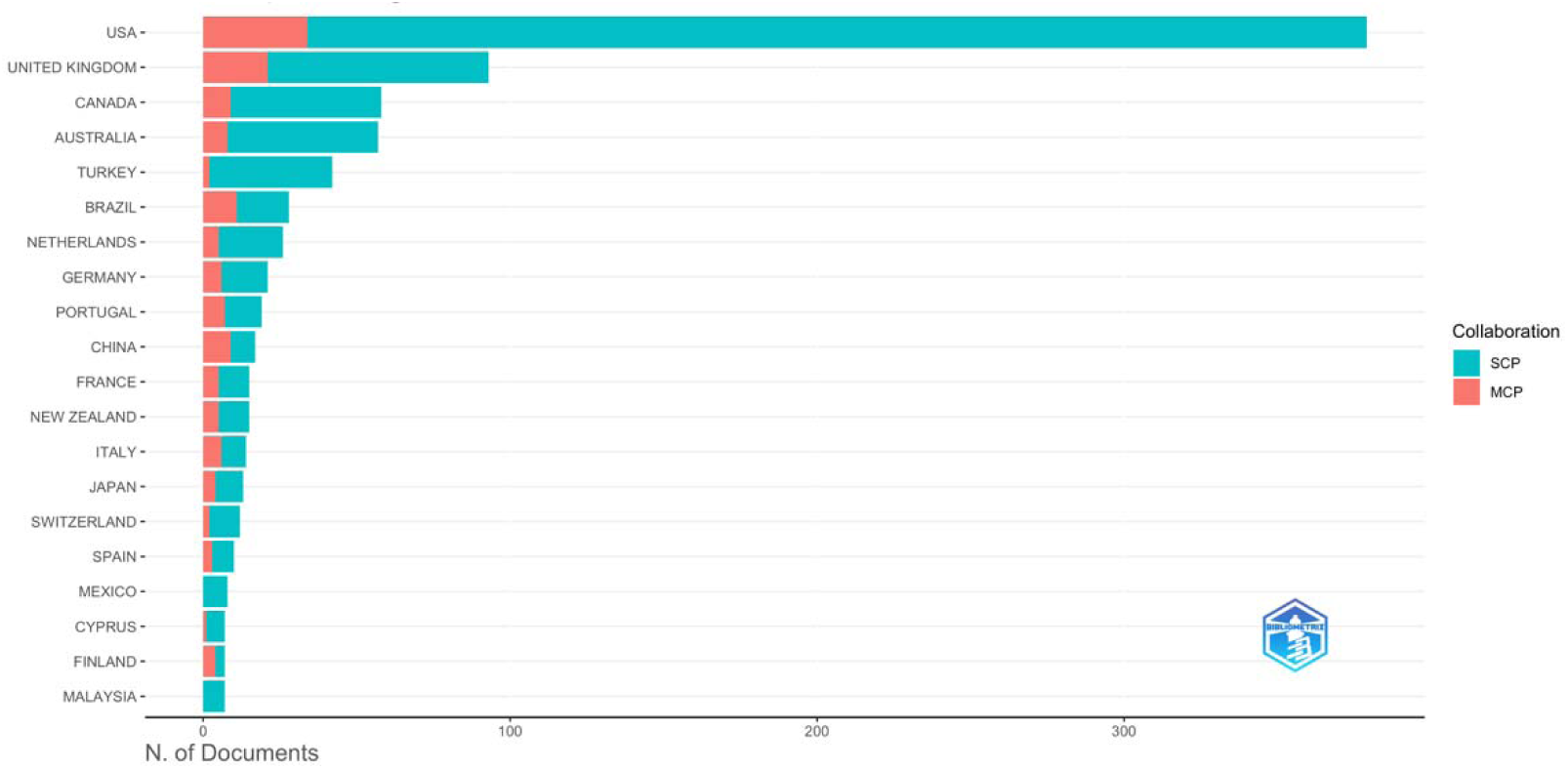
Number of publications and collaborations by corresponding author’s countries.

**Figure 2.**
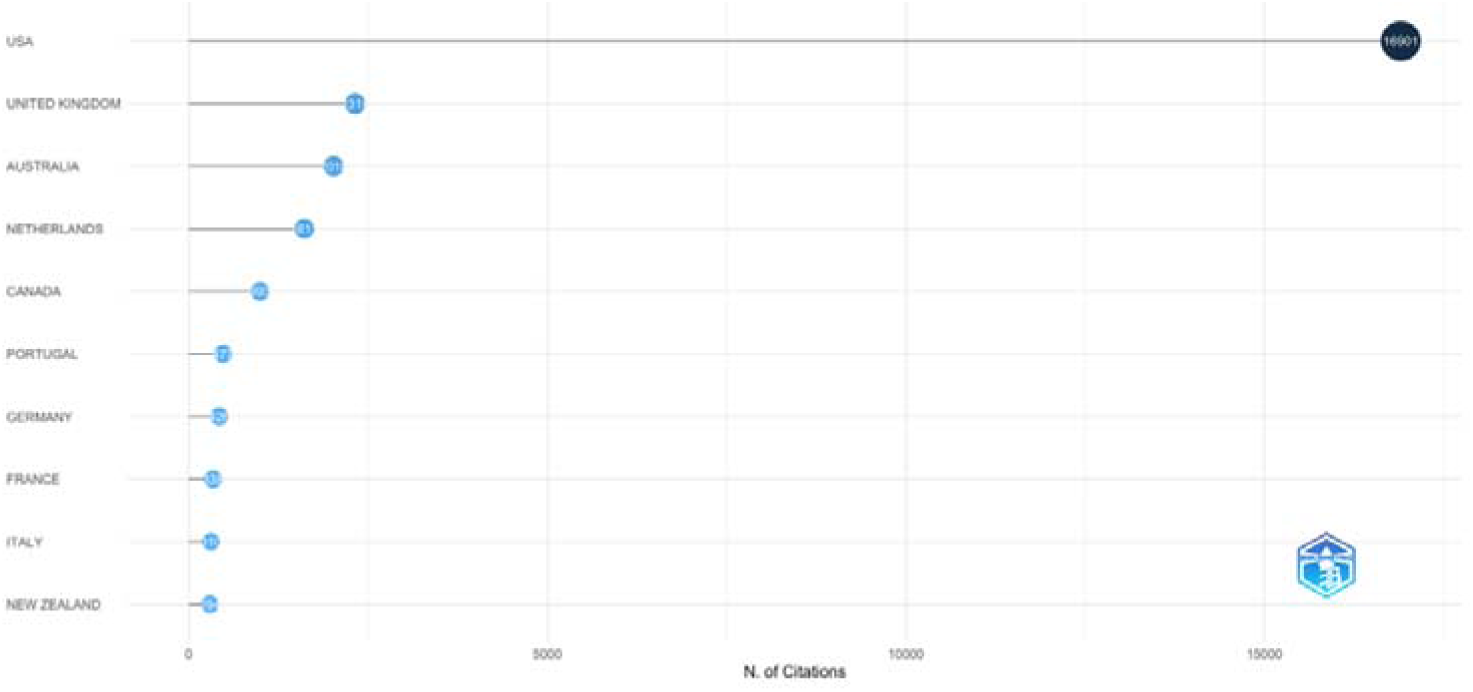
Top 10 most cited countries by corresponding author.

**Table 2.** Top countries by corresponding authors.

| Country | Articles | Articles % | SCP | MCP | MCP % |
| --- | --- | --- | --- | --- | --- |
| United States | 376 | 36.5 | 342 | 34 | 9 |
| United Kingdom | 93 | 9 | 72 | 21 | 22.6 |
| Canada | 58 | 5.6 | 49 | 9 | 15.5 |
| Australia | 57 | 5.5 | 49 | 8 | 14 |
| Turkey | 42 | 4.1 | 40 | 2 | 4.8 |
| Brazil | 28 | 2.7 | 17 | 11 | 39.3 |
| The Netherlands | 26 | 2.5 | 21 | 5 | 19.2 |
| Germany | 21 | 2 | 15 | 6 | 28.6 |
| Portugal | 19 | 1.8 | 12 | 7 | 36.8 |
| China | 17 | 1.7 | 8 | 9 | 52.9 |

Figure 3 further illustrates publication trends in ME and IE, with the U.S. showing a steep increase in ME and IE research paper production over the past two decades. This growth trajectory underscores its expanding research capacity and leadership in scientific contributions in these fields. Other countries like Canada, Brazil, Australia, and the U.K. display steady growth in research output, highlighting their sustained scholarship and collaboration.

**Figure 3.**
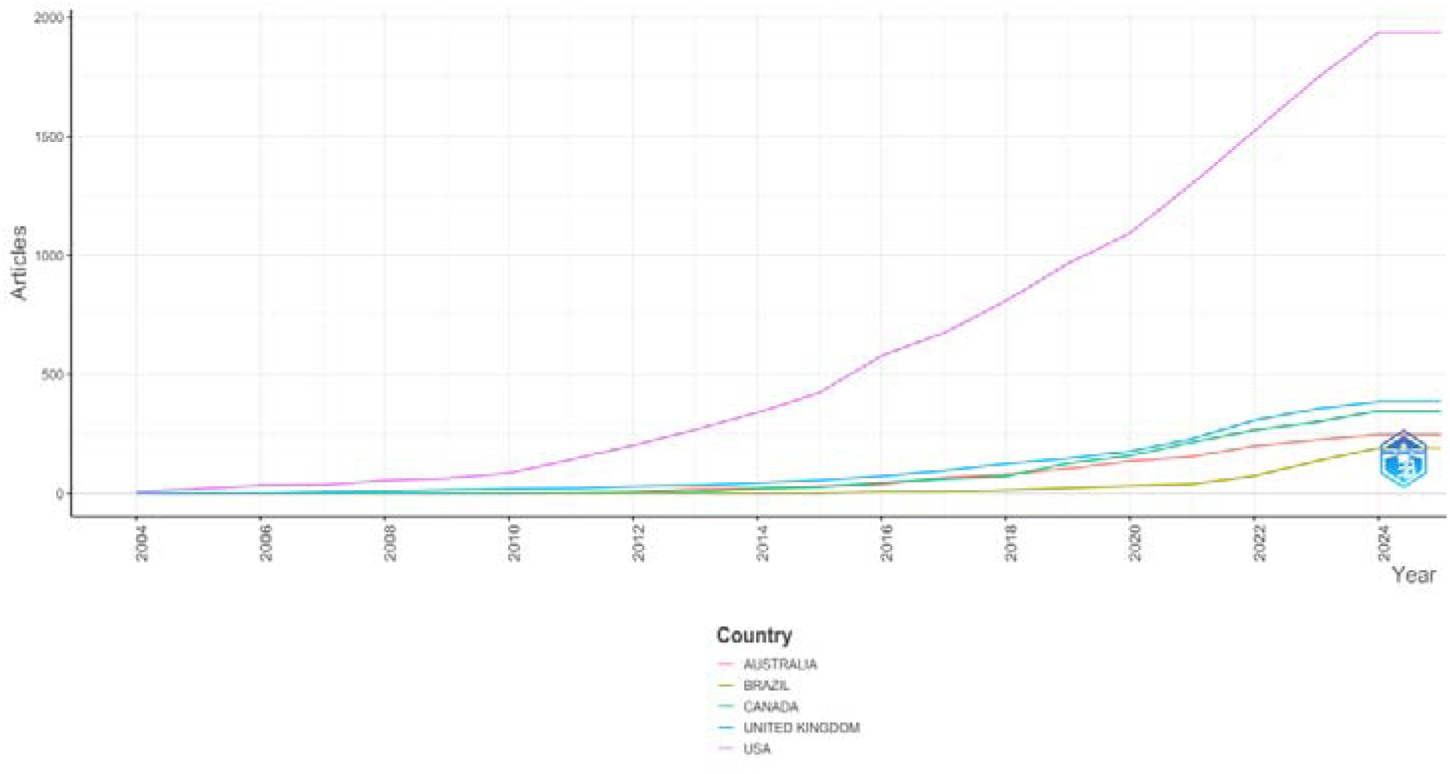
Top 5 countries article production over time.

### 3.2 Co-authorship analysis

Co-authorship analysis is a method of network analysis used to explore the collaboration patterns between authors of academic publications [32]. In this analysis, total link strength refers to the cumulative strength of all the connections in a given entity has with others in a network. It is a measure to quantify the intensity or frequency of collaborations [28]. Co-authorship using country as the unit of analysis examines the collaborative relationships between different countries based on shared authorship in academic publications. This analysis helps to identify international collaborations in ME and IE research and shows how these networks are distributed globally (Figure 4).

**Figure 4.**
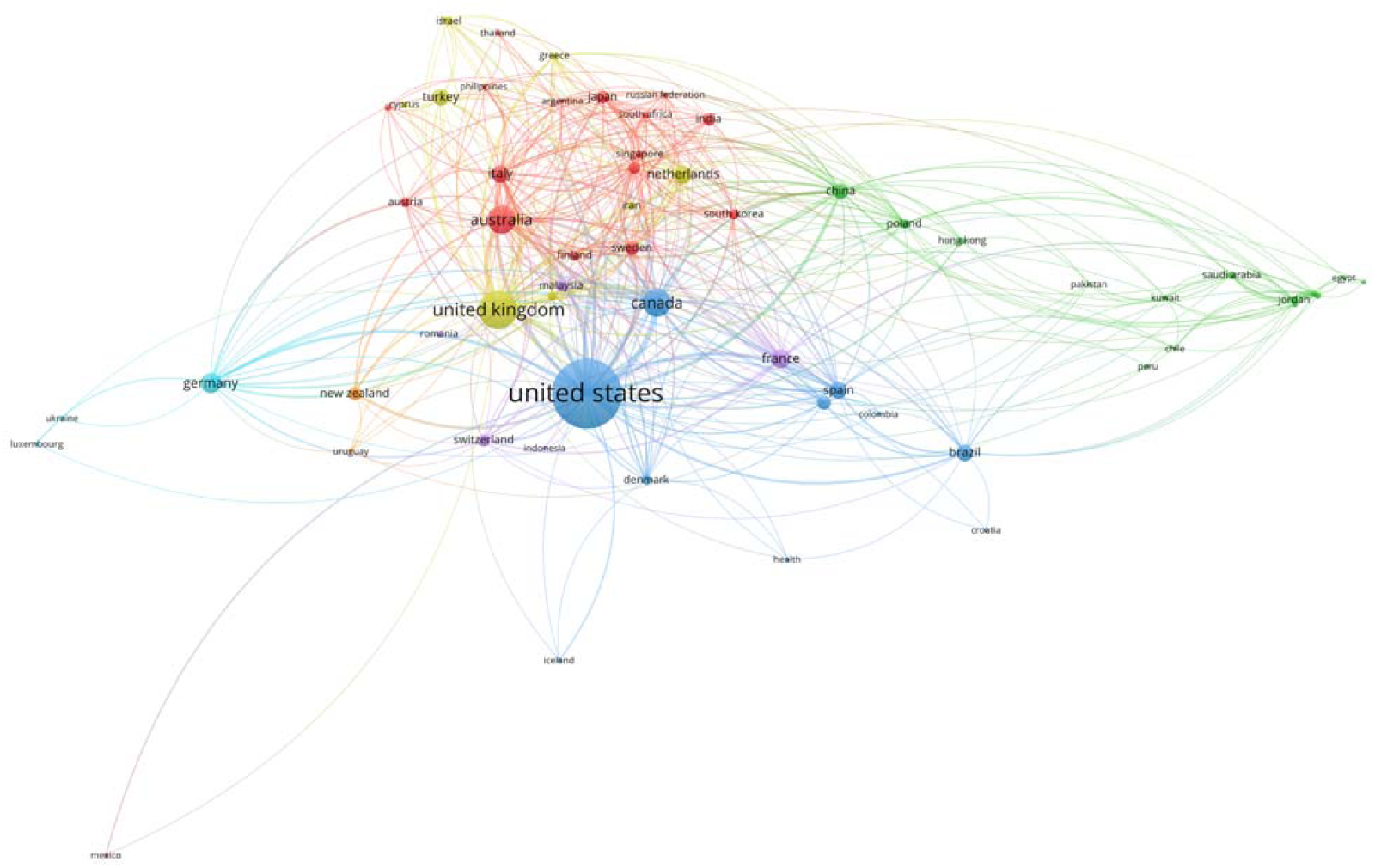
Countries with co-authorship links (5 or more publications)

Table 3 shows the top ten countries which play significant roles in international academic collaborations, including data on publication, citations, and total link strength. The U.S. has the highest total link strength of 141 (with 497 publications and 19,780 citations), indicating its central position in the global ME and IE research network and its substantial academic influence. The U.K. follows, with 131 publications and 4,558 citations, and a total link strength of 91. High income countries, such as Canada, Australia, France, Italy, Spain, Germany, and Japan are also prominent in the co-authorship network, highlighting their important roles in global research partnerships. China, as the only low- and middle- income country, also demonstrates active participation in international research in this field.

**Table 3.** Co-authorship analysis (Top 10 countries with highest total link strength)

| Rank | Country | Publications | Citations | Total link strength |
| --- | --- | --- | --- | --- |
| 1 | United States | 497 | 19780 | 141 |
| 2 | United Kingdom | 131 | 4558 | 91 |
| 3 | Canada | 88 | 1447 | 68 |
| 4 | Australia | 78 | 2518 | 53 |
| 5 | China | 23 | 189 | 43 |
| 6 | France | 26 | 578 | 40 |
| 7 | Italy | 24 | 483 | 40 |
| 8 | Spain | 19 | 340 | 36 |
| 9 | Germany | 40 | 871 | 31 |
| 10 | Japan | 17 | 129 | 24 |

### 3.3 Author analysis

In total, 3467 authors have contributed to ME or IE related research. The top ten most productive authors are shown in Table 4, with Mantzios, M. as the leading author having 26 publications and 494 citations, boasting a substantial total link strength of 51. Tylka, T., L. follows with 25 publications and 2,340 citations, with total link strength of 18 and Egan, H. is the third most prolific author with 21 publications, 252 citations and a total link strength of 48. Figure 6 highlights trends in research productivity among the top ten most productive authors over time which shows that authors such as Kristeller, J., Hecht, F. M. and Tylka, T. L. have maintained sustained publication outputs in ME and IE over the years, with a noticeable increase in publication activity in ME and IE-related research around 2017-2018 for several authors.

**Figure 5.**
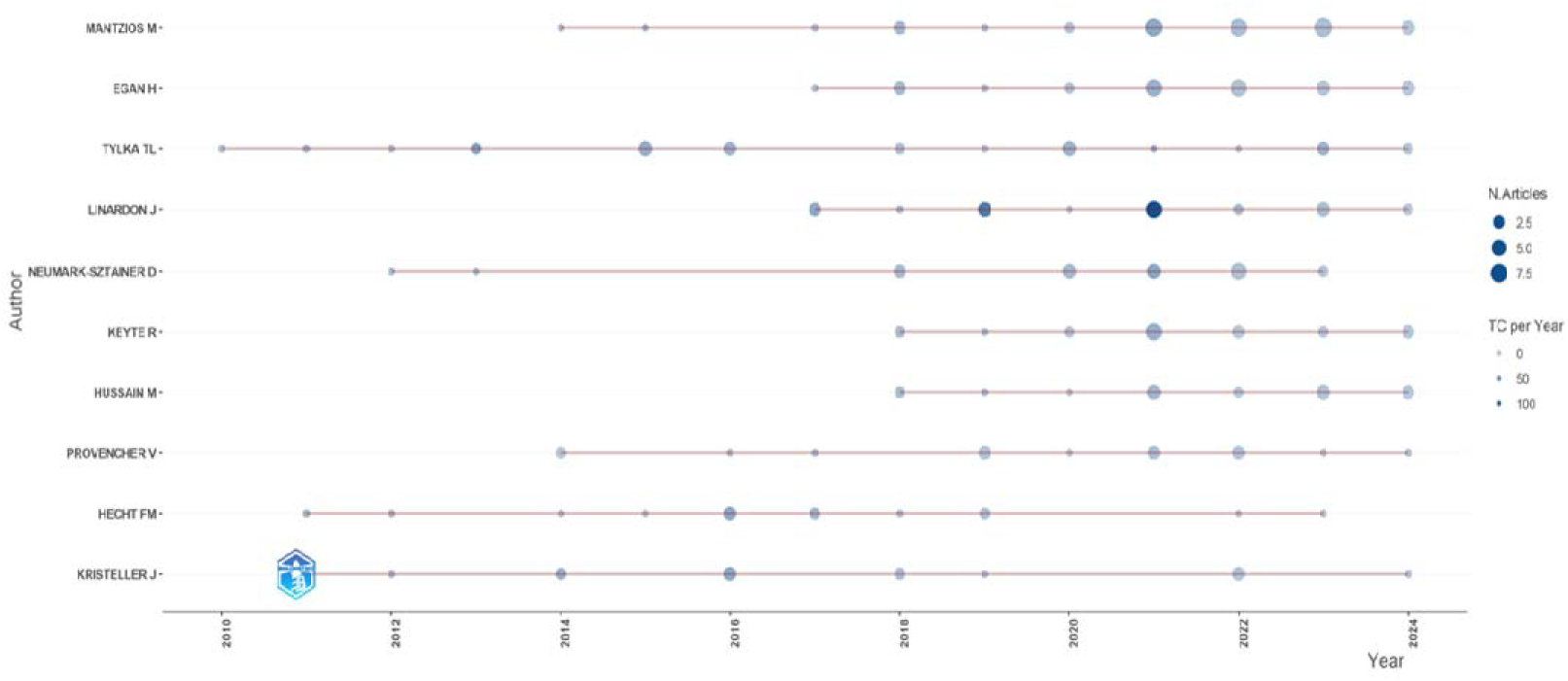
Top ten productive authors’ productivity over time (TC – total citations)

**Figure 6.**
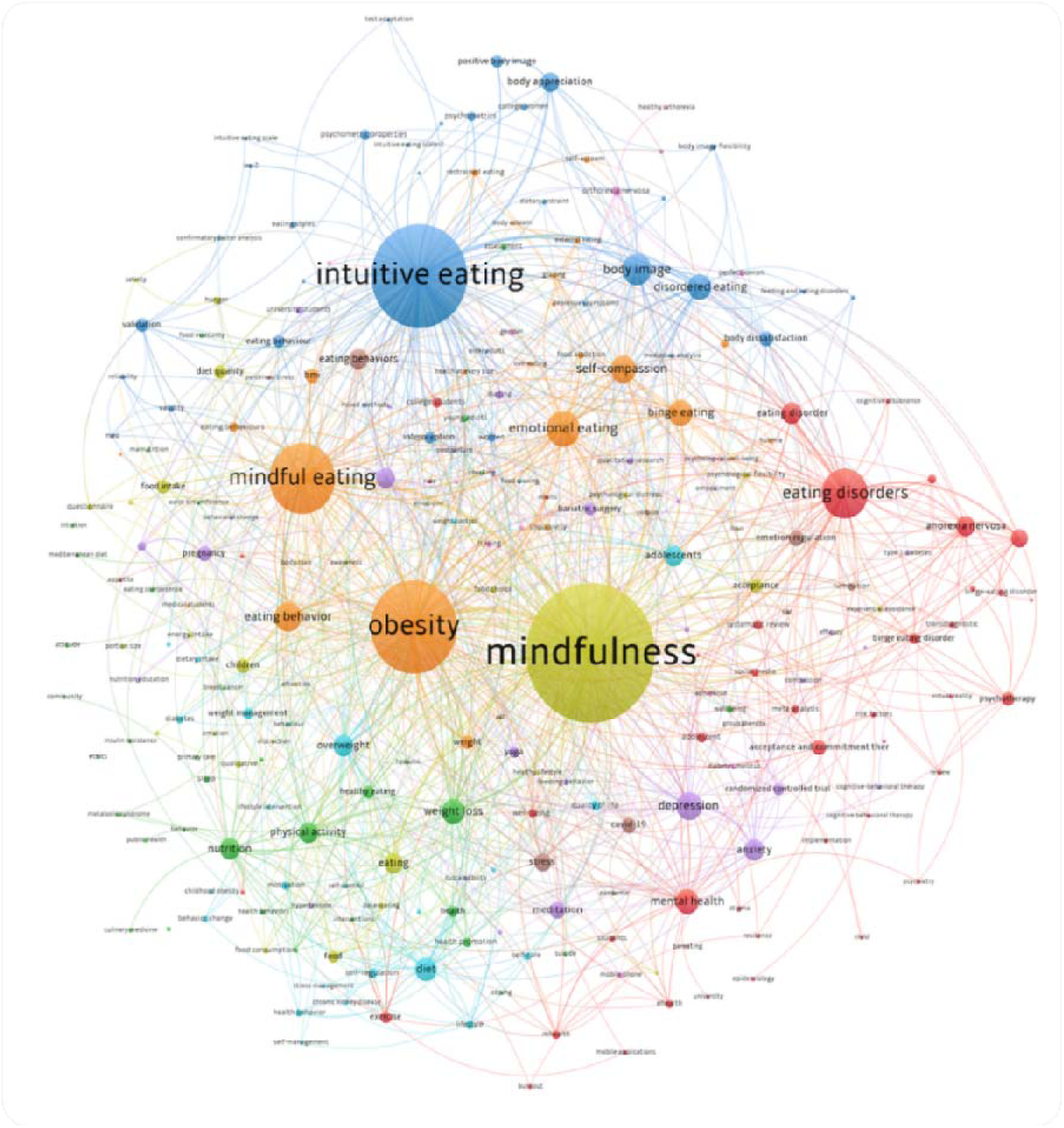
Network visualisation of author keywords (minimum occurrence of 5)

**Table 4.** Top ten most productive authors in ME and IE research.

| **Rank** | **Author name** | **Publications** | **Citations** | **Total link strength** |
| --- | --- | --- | --- | --- |
| 1 | Mantzios, M. | 26 | 494 | 51 |
| 2 | Tylka, T. L. | 25 | 2340 | 18 |
| 3 | Egan, H. | 21 | 252 | 48 |
| 4 | Neumark-Sztainer, D. | 16 | 424 | 26 |
| 5 | Keyte, R. | 13 | 175 | 34 |
| 6 | Linardon, J. | 14 | 587 | 19 |
| 7 | Provencher, V. | 13 | 219 | 20 |
| 8 | Hussain, M. | 13 | 156 | 38 |
| 9 | Kristeller, J. | 12 | 893 | 56 |
| 10 | Hecht, F. M. | 10 | 755 | 58 |

### 3.4 Citation analysis

In bibliometric analysis, the “*most cited documents*” and “*most cited references*” have different meanings. The “*most cited documents*” refer to papers that have been cited the most by other studies, reflecting the academic influence of these studies within the given field, helping to identify key research and trends in a specific area. On the other hand, the “*most cited references*” refer to references frequently cited within the analysed documents, showcasing the commonly cited research studies in the field [27]. A combined analysis of the two metrics helps to comprehensively understand the influence distribution and scientific knowledge base in a specific field. The top ten most cited articles on ME or IE research show a wide range of citation counts from 336 to 2,590. The highest citation count is 2,590, attributed to the paper titled “*The effect of mindfulness-based therapy on anxiety and depression: A meta-analytic review*” published in the Journal of Consulting and Clinical Psychology in 2010 (Table 5). Of the 1064 retrieved articles, a total of 43,196 references were cited. Table 6 presents the top ten most cited references in the dataset, showcasing the most influential research on ME and IE.

**Table 5.** Top ten cited articles in ME or IE research.

| Rank | Article Title | Year | Source | Citations |
| --- | --- | --- | --- | --- |
| 1 | The effect of mindfulness-based therapy on anxiety and depression: A meta-analytic review | 2010 | Journal of Consulting and Clinical Psychology | 2590 |
| 2 | Meditation programs for psychological stress and well-being | 2014 | JAMA Intern Medicine | 1460 |
| 3 | The unhealthy = tasty intuition and its effects on taste inferences, enjoyment, and choice of food products | 2006 | Journal of Marketing | 908 |
| 4 | Diversity in the determinants of food choice: A psychological perspective | 2009 | Food quality and preference | 662 |
| 5 | Self-enhancement: food for thought | 2008 | Perspectives on Psychological Science | 563 |
| 6 | The intuitive eating scale–2: Item refinement and psychometric evaluation with college women and men | 2013 | Journal of counseling psychology | 462 |
| 7 | Development and validation of a self-report measure of mentalizing: The reflective functioning questionnaire | 2016 | PLOS One | 441 |
| 8 | Mindfulness-based eating awareness training for treating binge eating Disorder: the conceptual foundation | 2014 | Eating Disorder and Mindfulness | 376 |
| 9 | Development and psychometric evaluation of a measure of intuitive eating | 2006 | Journal of Counseling Psychology | 366 |
| 10 | Mindfulness-based interventions for obesity-related eating behaviours: a literature review | 2014 | Obesity Reviews | 336 |

**Table 6.** Top 10 cited references in ME or IE research.

| Rank | Article Title | Year | Source | Citations |
| --- | --- | --- | --- | --- |
| 1 | A systematic review of the psychosocial correlates of intuitive eating among adult women | 2016 | Appetite | 76 |
| 2 | Development and psychometric evaluation of a measure of intuitive eating | 2006 | Journal of Counseling Psychology | 58 |
| 3 | The Body Appreciation Scale-2: item refinement and psychometric evaluation | 2015 | Body Image | 51 |
| 4 | Does mindfulness matter? Everyday mindfulness, mindful eating and self-reported serving size of energy dense foods among a sample of South Australian adults | 2013 | Appetite | 49 |
| 5 | Statistical Power Analysis for the Behavioral Sciences | 1988 | Taylor & Francis | 48 |
| 6 | Is intuitive eating the same as flexible dietary control? Their links to each other and well-being could provide an answer | 2015 | Appetite | 46 |
| 7 | Using self-report assessment methods to explore facets of mindfulness | 2006 | Assessment | 44 |
| 8 | A structured literature review on the role of mindfulness, mindful eating and intuitive eating in changing eating behaviours: effectiveness and associated potential mechanisms | 2017 | Nutrition research reviews | 39 |
| 9 | Diagnostic and statistical manual of mental disorders 5: A quick glance | 2013 | Indian Journal of Psychiatry | 38 |
| 10 | Development and validation of the mindful eating questionnaire. | 2009 | Journal of the American Dietetic Association | 38 |

### 3.5 Keywords analysis

#### 3.5.1 Cluster analysis

We used VOSviewer to conduct the co-occurrence analysis using author keyword as the unit of analysis as they typically reflect the research theme of the article more directly, whereas index keywords are often provided by databases and tend to focus on broader subject classifications which may not accurately capture the specific research topics [33]. There were 3,647 author keywords across all publications and the analysis shows the most commonly occurring keywords (minimum occurrence of 5) in the fields of ME and IE research (Figure 7). These are grouped into 9 clusters in the figure, with each dot representing an author keyword and the size of dots reflects the frequency of occurrences in the dataset. Among the top three coloured clusters, the red cluster has 45 keywords, 119 occurrences, and 98 links, with a total link strength of 298. Within this cluster, the top keywords associated with eating disorders are mental health, anorexia nervosa, binge eating, and therapies such as acceptance and commitment therapy and psychotherapy. The green cluster has 35 keywords, 51 occurrences, and 74 links, with total link strength of 189. Within this cluster, the top keywords associated with weight loss are physical activity, nutrition, and healthy eating. The blue cluster has 35 keywords, 296 occurrences, and 158 links, and with total link strength of 705. Within this cluster, the top keywords associated with intuitive eating are body image, body dissatisfaction, body appreciation, and positive body image. The most frequent author keyword co-occurrences include ‘mindfulness’, ‘intuitive eating’, and ‘obesity’ (Table 7).

**Figure 7.**
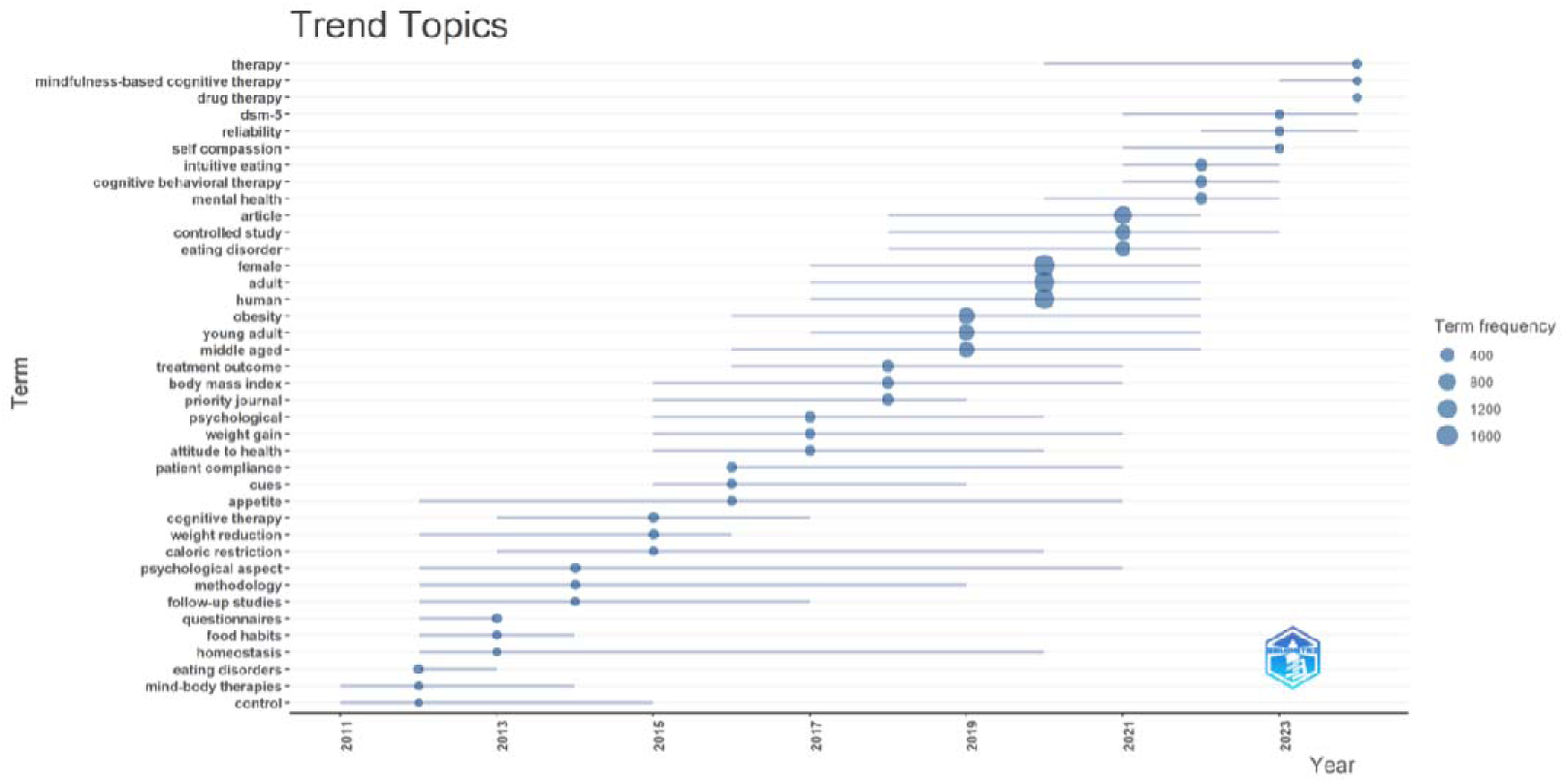
Trend topics in ME and IE (generated by Bibliometrix)

**Table 7.** Top ten author keyword co-occurrence.

| **Rank** | **Author keyword** | **Occurrences** | **Total link strength** |
| --- | --- | --- | --- |
| 1 | mindfulness | 429 | 1185 |
| 2 | intuitive eating | 296 | 714 |
| 3 | obesity | 258 | 816 |
| 4 | mindful eating | 184 | 475 |
| 5 | eating disorders | 119 | 313 |
| 6 | emotional eating | 80 | 251 |
| 7 | body image | 68 | 209 |
| 8 | eating behavior | 62 | 182 |
| 9 | depression | 59 | 197 |
| 10 | self-compassion | 57 | 157 |

#### 3.6.2 Trend analysis

We also used Bibliometrix to analyse research trends related to ME and IE in the past decade. As shown in Figure 7, the evolution of topics such as ‘*mindfulness-based cognitive therapy*’ and ‘*drug therapy*’ which have become popular in recent years. Core issues like ‘*homeostasis*’, ‘*appetite*’, and ‘*psychological aspects*’ have sustained attention throughout the entire timeframe, indicating their long-term significance. Emerging areas of focus include ‘*intuitive eating*’ and ‘*self-compassion’* which have gained more prominence in recent years. The research also reflects diversity in subjects, covering various age groups including ‘*young adult*’ and ‘*middle-aged*’ adults and genders.

## 4. DISCUSSION

### 4.1 Principal findings

This study provides the most updated analysis on ME and IE research over the last two decades. Trend analysis shows that psychological aspects, assessment tools, research methodology and methods, have gained increasing attention, reflecting the dynamic progression on ME and IE research. The cluster analysis reveals nine clusters in ME and IE research, with emerging areas like ‘*intuitive eating*’, ‘*mindful eating*’, and ‘*mindfulness*’ gaining prominence over time. It also uncovers commonly used keywords related to eating disorders, weight loss, and obesity showing a trend towards clinical and psychological interventions that involve IE and ME rather than alternatives such as public health initiatives.

Productivity and co-authorship analysis results on countries, authors, and institutions highlights the U.S. as the dominant force in global research on ME and IE, leading in both publication and citation counts, as well as extensive international collaborations. The UK and Australia also play significant roles, reflecting strong participation in academic networks. Notably, European countries consistently rank high in research output and citations with emerging nations, particularly China and Turkey, increasing their influence in global research on this topic. The analysis of co-authorship reveals robust collaboration patterns, with the U.S. showing the highest total link strength, followed by the U.K. and Canada. Although mindfulness developed from Buddhist traditions in Eastern philosophy, most research in this review originated in Western countries which have yet to adopt mindful or intuitive eating practices in their eating cultures. Hence, more multi-cultural collaboration could enhance research by not only improving the development of ME and IE interventions but also outcome measures. A significant number of authors contribute to research in ME and IE, with several key researchers maintaining consistent publication activity, particularly in recent years. The most influential articles in the fields of ME and IE were also identified, which can help researchers identify research hotspots and relevant concepts and theories.

### 4.2 Future implications

This bibliometric analysis of ME and IE research over the past two decades provides valuable insights for future research directions. First, the dominance of countries like the U.S., U.K., and Australia highlights the importance of fostering international collaborations, especially with emerging nations which are increasingly contributing to global research. Expanding partnerships between developed and developing countries could diversify research perspectives and lead to advances in scientific research in ME and IE in particular in the global south [34]. Future research efforts should prioritise building global networks to foster knowledge exchange between different nations and institutions, which could potentially elevate research impact in these fields on a worldwide scale.

Second, keyword analysis reveals important emerging areas such as ‘*mindfulness*’, ‘*mindful eating*’, and ‘*intuitive eating*’, along with growing attention to psychological aspects of IE and ME. Future studies could explore the neuronal mechanisms and expand these topics further to highlight the role of interoception, decentering, acceptance and self-compassion to increase autonomy and motivation and reduce shame (Tapper, 2022). The increasing diversity of research subjects, methodologies, and methods in this research area also suggests that it is important to strengthen interdisciplinary research and integrate insights from diverse disciplines, such as psychology, neurobiology, nutrition, public health and policy etc. By addressing these gaps and expanding international and interdisciplinary collaborations, researchers could strengthen the overall impact of ME and IE research in academic and real-world settings.

### 4.3 Strengths and limitations

A key strength of this study lies in the application of comprehensive bibliometric software including Bibliometrix and VOSviewer which effectively captured the overall landscape of ME and IE research, illustrating key trends in these fields. However, there are some limitations to consider. First, only articles and reviews published in English were included in the analysis, excluding grey literature and research published in other languages, which may have led to the omission of relevant trends. Second, while Scopus was chosen as the primary database, using other databases like Web of Science or PubMed could potentially produce different results due to differences in indexing and coverage. Third, the information extracted by bibliometric analysis software cannot guarantee 100% accuracy, such as authors names. This may be because authors’ names can appear in different forms across various articles, leading to dispersed data. Although the authors of this review manually cleaned and standardized the data before undertaking the analysis, inaccuracies may still occur. Lastly, although this bibliometric analysis provides useful insights into publication trends and metrics, it does not assess the quality of the included studies.

## 5. CONCLUSION

In this review, valuable insights into the research landscape of ME and IE are gained through comprehensive analyses of countries, institutions, authors, citations, journals, and keywords. By examining collaboration networks between countries, institutions, and authors, as well as identifying emerging trends through keyword analysis, the study highlights the current state of research in these fields. The results offer a clear understanding of the research dynamics in the past two decades, enabling researchers to identify key trends and potential collaborations for future studies on ME and IE.

## Data Availability

All data produced in the present study are available upon reasonable request to the authors.

## ABBREVIATIONS

MCP: Collaboration within countries
SCP: Collaboration between countries
IE: Intuitive eating
ME: Mindful eating
U.S.: United States of America
U.K.: United Kingdom

## Acknowledgements

This work was funded by The Burdett Trust for Nursing via a grant awarded in 2022.

## Author contributions

Siobhan O’Connor and Mengying Zhang designed and conducted the study, collated and analysed the data, and Mengying Zhang drafted the manuscript. Angeliki Bogossian, Emma Stanmore and Trudi Edginton assisted in the visualization of the bibliometric analysis, provided research expertise, and edited the manuscript. As this work’s guarantor, Siobhan O’Connor has full access to all study dataset and is responsible for the data’s integrity and accuracy.

## REFERENCES

1. Cifuentes L, Acosta A. Homeostatic regulation of food intake. Clinics and research in hepatology and gastroenterology. 2022;46(2):101794.

2. Reichenberger J, Schnepper R, Arend AK, Richard A, Voderholzer U, Naab S, et al. Emotional eating across different eating disorders and the role of body mass, restriction, and binge eating. Int J Eating Disord. 2021;54(5):773–84.

3. Pagliai G, Dinu M, Madarena MP, Bonaccio M, Iacoviello L, Sofi F. Consumption of ultra-processed foods and health status: a systematic review and meta-analysis. British journal of nutrition. 2021;125(3):308–18. doi: 10.1017/S0007114520002688.

4. WHO. Obesity and overweight 2024. Available from: https://www.who.int/news-room/fact-sheets/detail/obesity-and-overweight.

5. Chooi YC, Ding C, Magkos F. The epidemiology of obesity. Metabolism. 2019;92:6–10. doi: 10.1016/j.metabol.2018.09.005.

6. Safaei M, Sundararajan EA, Driss M, Boulila W, Shapi’i A. A systematic literature review on obesity: Understanding the causes & consequences of obesity and reviewing various machine learning approaches used to predict obesity. Comput Biol Med. 2021;136:104754-. doi: 10.1016/j.compbiomed.2021.104754.

7. Brinkworth GD, Buckley JD, Noakes M, Clifton PM, Wilson CJ. Long-term Effects of a Very Low-Carbohydrate Diet and a Low-Fat Diet on Mood and Cognitive Function. Archives of Internal Medicine. 2009;169(20):1873–80. doi: 10.1001/archinternmed.2009.329.

8. Daneshzad E, Keshavarz SA, Qorbani M, Larijani B, Azadbakht L. Association between a low-carbohydrate diet and sleep status, depression, anxiety, and stress score. J Sci Food Agric. 2020;100(7):2946–52. doi: 10.1002/jsfa.10322.

9. O’Neill B, Raggi P. The ketogenic diet: Pros and cons. Atherosclerosis. 2020;292:119–26.

10. Watanabe M, Tuccinardi D, Ernesti I, Basciani S, Mariani S, Genco A, et al. Scientific evidence underlying contraindications to the ketogenic diet: An update. Obes Rev. 2020;21(10):e13053-n/a. doi: 10.1111/obr.13053.

11. Mantzios M. (Re)defining mindful eating into mindful eating behaviour to advance scientific enquiry. Nutr Health. 2021;27(4):367–71. doi: 10.1177/0260106020984091. PubMed Central PMCID: PMC33356846.

12. Gilbert P. Introducing compassion-focused therapy. Advances in psychiatric treatment : the Royal College of Psychiatrists’ journal of continuing professional development. 2009;15(3):199–208. doi: 10.1192/apt.bp.107.005264.

13. Warren JM, Smith N, Ashwell M. A structured literature review on the role of mindfulness, mindful eating and intuitive eating in changing eating behaviours: Effectiveness and associated potential mechanisms. Nut Res Rev. 2017;30(2):272–83. doi: 10.1017/S0954422417000154. PubMed Central PMCID: PMC28718396.

14. Tylka TL, Wilcox JA. Are intuitive eating and eating disorder symptomatology opposite poles of the same construct? J Couns Psychol. 2006;53(4):474–85. doi: 10.1037/0022-0167.53.4.474.

15. Linardon J, Tylka TL, Fuller-Tyszkiewicz M. Intuitive eating and its psychological correlates: A meta-analysis. The International journal of eating disorders. 2021;54(7):1073–98. doi: 10.1002/eat.23509.

16. Carrière K, Khoury B, Günak MM, Knäuper B. Mindfulness-based interventions for weight loss: a systematic review and meta-analysis. Obes Rev. 2018;19(2):164–77. doi: 10.1111/obr.12623.

17. Robinson L, Gordon G, Werthmann J, Campbell IC, Schmidt U. The outcomes of mindfulness-based interventions for Obesity and Binge Eating Disorder: A meta-analysis of randomised controlled trials. Appetite. 2021;166. doi: 10.1016/j.appet.2021.105464. PubMed Central PMCID: PMC34146647.

18. Grider HS, Douglas SM, Raynor HA. The Influence of Mindful Eating and/or Intuitive Eating Approaches on Dietary Intake: A Systematic Review. Journal of the Academy of Nutrition and Dietetics. 2021;121(4):709–27.e1. doi: 10.1016/j.jand.2020.10.019. PubMed Central PMCID: PMC33279464.

19. Donthu N, Kumar S, Mukherjee D, Pandey N, Lim WM. How to conduct a bibliometric analysis: An overview and guidelines. Journal of business research. 2021;133:285–96.

20. Yuan K, Zhang X, Wu B, Zeng R, Hu R, Wang C. Research trends between diabetes mellitus and bariatric surgery researches: Bibliometric analysis and visualization from 1998 to 2023. Obes Rev. 2024;25(6):e13730-n/a. doi: 10.1111/obr.13730.

21. Khan A, Choudhury N, Uddin S, Hossain L, Baur LA. Longitudinal trends in global obesity research and collaboration: a review using bibliometric metadata. Obes Rev. 2016;17(4):377–85. doi: 10.1111/obr.12372.

22. Petermann-Rocha F, Diaz-Toro F, Valera-Gran D, Navarrete-Muñoz EM. Bibliometric analysis of research on sarcopenic obesity: a review of scientific literature. Obes Rev. 2024;25(9):e13784-n/a. doi: 10.1111/obr.13784.

23. Burnham JF. Scopus database: a review. Biomedical digital libraries. 2006;3:1–8.

24. Montoya FG, Alcayde A, Baños R, Manzano-Agugliaro F. A fast method for identifying worldwide scientific collaborations using the Scopus database. Telematics and informatics. 2018;35(1):168–85. doi: 10.1016/j.tele.2017.10.010.

25. Aletaha A, Soltani A, Dokhani F. Evaluating obesity publications: from bibliometrics to altmetrics. Journal of diabetes and metabolic disorders. 2021;20(1):391–405. doi: 10.1007/s40200-021-00758-7.

26. Corrêa EL, Cotian LFP, Lourenço JW, Lopes CM, Carvalho DR, Strobel R, et al. Overview of the Last 71 Years of Metabolic and Bariatric Surgery: Content Analysis and Meta-analysis to Investigate the Topic and Scientific Evolution. Obes Surg. 2024;34(5):1885–908. doi: 10.1007/s11695-024-07165-w.

27. Aria M, Cuccurullo C. bibliometrix: An R-tool for comprehensive science mapping analysis. Journal of informetrics. 2017;11(4):959–75.

28. Van Eck NJ, Waltman L. VOSviewer manual. Manual for VOSviewer version. 2011;1(0).

29. Roth B, Creaser T. Mindfulness meditation-based stress reduction: experience with a bilingual inner-city program. Nurse Pract. 1997;22(3):150–2,4,7passim.

30. Gast J, Hawks SR. Weight Loss Education: The Challenge of a New Paradigm. Health Educ Behav. 1998;25(4):464–73. doi: 10.1177/109019819802500405.

31. Bornmann L, Daniel H-D. What do citation counts measure? A review of studies on citing behavior. J Doc. 2008;64(1):45–80. doi: 10.1108/00220410810844150.

32. De Stefano D, Giordano G, Vitale MP. Issues in the analysis of co-authorship networks. Quality & Quantity. 2011;45:1091–107.

33. University of Southern California. Searching Solutions: Keywords vs Indexed 2025 [cited 2025 January 14th]. Available from: https://libguides.usc.edu/c.php?g=631331&p=4459632.

34. Reidpath DD, Allotey P. The problem of ‘trickle-down science’from the Global North to the Global South. BMJ Global Health. 2019;4(4):e001719.

